# Mortality and use of angiotensin converting enzyme inhibitors in Covid 19 disease – a systematic review

**DOI:** 10.1101/2020.05.29.20116483

**Authors:** José Pedro L. Nunes

## Abstract

**Background:** Interest exits concerning the use of angiotensin converting enzyme inhibitors (ACEi) in patients with Covid-19 disease.

**Objectives:** To perform a systematic review on mortality associated to the use of ACEi in patients with Covid 19 disease.

**Methods:** Search in Medline (PubMed), in ISI Web of Knowledge and in medRxiv database; use of other sources.

**Results:** A total of 33 articles were evaluated. Concerning the papers used to produce the meta-analyses, seven studies were selected, five of which were used. These five studies involved a total number of 944 patients treated with ACEi and 5173 not treated with ACEi. Increased mortality was seen in association to the use of ACEi in the context of Covid-19 disease (ACEi users versus non-users; odds ratio, 1.48; 95% confidence interval [CI], 1.02 to 2.15; P=0.04). When compared to mortality in patients treated with angiotensin receptor blockers, mortality of patients treated with ACEi was not significantly different (odds ratio, 0.96; 95% confidence interval [CI], 0.76 to 1.21; P=0.74). Concerning the remaining reports, different types of data adjustments were used by several authors, after which increased mortality was not seen in association to the use of ACEi in this context.

**Conclusions:** ACEi use could act as a marker of increased mortality risk in some but not all Covid-19 disease settings. The data now presented do not prove a causal relation but argue in favor of carrying out clinical trials studying ACEi in Covid-19 patients, in order to establish the safety of ACEi use in this context.

## Introduction

An epidemic of viral disease caused by a new Coronavirus, Sars-Cov-2, is currently underway in most regions of the world. There is interest concerning angiotensin converting enzyme inhibitors (ACEi) use in this context, since the virus appears to interact with the angiotensin converting enzyme type 2 (ACE2) ^1^.

Although ACEi and angiotensin receptor blockers are sometimes evaluated together, they do not have a common mechanism of action, and therefore a separate evaluation of ACEi use in this context may be of interest. In the present report, a systematic review was carried out, looking at published reports studying the association between ACEi use and mortality in patients with Covid-19, the disease caused by the new Coronavirus. The aim of the study was to use currently available data to tentatively evaluate if a relation exists between ACEi use and patient mortality in this context.

## Methods

### Search strategy

The study started with a search on Medline (PubMed), in ISI Web of Knowledge and in medRxiv databases, using the query “Covid-19” AND “ACE inhibitor” AND “mortality” (first query) and “Covid-19” AND “angiotensin converting enzyme inhibitor” AND “mortality” (second query). The search took place on June 16-21, 2020, and no articles were excluded based on publication date. The queries resulted in different sets of articles being found, as presented in Figure 1, prepared in accordance with the Preferred Reporting Items for Systematic Reviews and Meta-Analyses statement. Further additional studies were identified in other relevant sources, including the sites of major medical journals (Figure 1).

**Figure 1.**
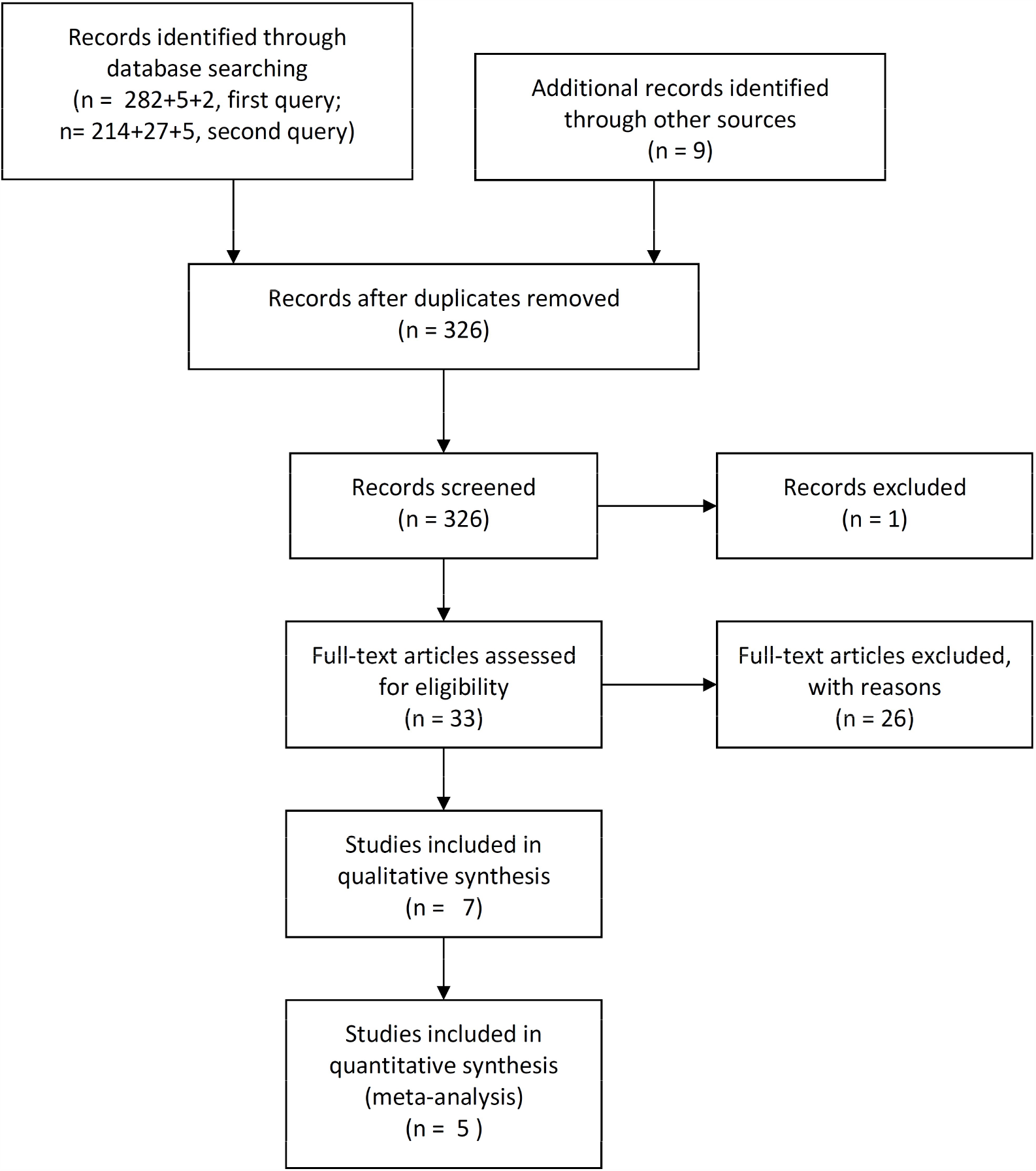
Flow diagram of studies selection.

### Inclusion criteria

Only human studies with original data were included.

### Exclusion criteria

Excluded were: mechanistic studies; animal studies; case reports; editorials; review papers; study protocols; duplicate studies, if found; systematic reviews and/or meta-analyses; guidelines; genetic or pathological studies.

### Statistical analysis

Meta-analysis was carried out by using the Comprehensive Meta-analysis Software, V.2.0 (Biostat, New Jersey, USA). Fixed effects or random effects analyses were carried out, depending on the degree of heterogeneity of the data (fixed effects for I squared values <50; random effects otherwise). Mortality was the only parameter under study, and the odds ratio was calculated. A level of significance of 0.05 was used.

### Quality assessment of studies and data extraction

Global article quality assessment was carried out according to the method used by Haffar *et a*l., concerning the articles used for the meta-analyses ^2^.

## Results

A total of 33 articles were identified and selected for further study (listed in Supplementary Table 1).

**Table 1.** Major data from the selected papers. ACEi – angiotensin converting enzyme inhibitors. ARB – angiotensin receptor blockers. For references see text.

| Authors | Population | ACEi users | Non ACEi users | ARB users |
| --- | --- | --- | --- | --- |
| Li et al. | Observational single-center case series of the 1178 hospitalized patients with COVID-19 infections at the Central Hospital of Wuhan, China, from January 15 to March 15, 2020. Data reported for 362 patients with arterial hypertension. Median age 55.5 years (interquartile range, 38-67 years). Overall mortality: 11.03% (21.27% for patients with arterial hypertension). Criterion: use of drugs at the time of admission that continued through hospitalization. | 35 ACEi users.<br>28 survivors;<br>7 non-survivors. | 327 non-ACEi users.<br>257 survivors;<br>70 non-survivors. | 83 ARB users.<br>68 survivors;<br>15 non-survivors. |
| Richardson et al. | Observational case series of 5700 patients with COVID-19 admitted to 12 hospitals in New York. Median age 63 years, (interquartile range 52-75 years). Data reported for patients with arterial hypertension. Overall mortality: 20.99% (28.11% for patients with arterial hypertension). Criterion: home medication at admission. | 168 ACEi users.<br>113 patients discharged;<br>55 non-survivors. | 1198 Non-ACEi users.<br>869 patients discharged;<br>329 non survivors. | 245 ARB users.<br>170 patients discharged;<br>75 non-survivors. |
| Rossi et al. | Observational study of all 2653 symptomatic patients who tested positive for SARS-CoV-2 from February 27 to April 2, 2020 in the province of Reggio Emilia. Mean age 63.2 years. Overall mortality: 8.18%. Criterion: use of drugs in previous year. | 450 ACEi users.<br>56 non-survivors. | 2203 Non-ACEi users.<br>161 non-survivors. | 368 ARB users.<br>52 non-survivors. |
| Jung et al. | Nationwide cohort study using the Korean Health Insurance Review and Assessment database: 5179 | 45 ACEi users.<br>0 non-survivors. | 5134 non-ACEi users.<br>84 non-survivors. | 732 ARB users.<br>33 non-survivors |
|  | patients with Covid-19; 1954 patients hospitalized.<br>Mean age: 44.6 years overall cohort, 62.5 years patients using either ARB or ACEi.<br>Overall in-hospital mortality: 4.3%.<br>Criterion: drug use at 1-30 days before the index date. |  |  |  |
| Felice *et al.* | Single center study of 133 hypertensive subjects with Covid-19 disease in March 2020.<br>Mean age 73.1 years for ACEi users, 69.0 years for ARB users.<br>Overall mortality: 24.8%.<br>Criterion: chronic use of drugs. | 40 ACEi users.<br>8 non-survivors. | 93 non-ACEi users.<br>25 non-survivors. | 42 ARB users.<br>7 non-survivors. |
| Meng *et al.* | Observational study of 476 patients recruited from January 1 to February 15, 2020 at three hospitals in Wuhan, Shanghai and Anhui.<br>Median age of analyzed subjects 64.5 years (interquartile range, 55.8–69.0 years). Data from 42 patients receiving antihypertensive therapy.<br>Overall mortality: 2.38%. | 2 ACEi users.<br>0 non-survivors. | 40 non-ACEi users.<br>1 non-survivor. | 14 ARB users.<br>0 non-survivors. |
| Bravi *et al.* | Observational study of 1603 adults with SARS-CoV-2 infection from two Italian provinces.<br>Mean age of 58.0 years.<br>Overall mortality: 9.6%.<br>Criterion: background pharmacological treatment up to the previous two years (from prescription database), integrated with clinical chart information for hospitalized subjects. | 251 ACEi users.<br>45 non-survivors. | 1352 non-ACEi users.<br>109 non-survivors. | 228 ARB users.<br>46 non-survivors. |

### Reports under meta-analyses

A number of articles failed to include the data of interest for the present purpose - and this was the major reason to exclude reports from entering meta-analyses. The precise number of deaths or specific information on ACEi users were among the data not presented in some reports. One article, initially selected, was retracted by the authors on June 4, 2020, and was therefore excluded, leaving a total of seven selected studies. Two of the selected reports (presented in Table 1) had no fatalities in one of the groups, making these data unsuitable to be used by the meta-analysis software ^3,4^. Five studies were used to produce the meta-analyses ^5–9^ (Figures 2 and 3). These five studies involved a total number of 944 patients treated with ACEi and 5173 not treated with ACEi. The main results concerning ACEi use and mortality are presented in Table 1. Some reports presented data concerning only hypertensive patients, whereas others did not (Table 1). Increased mortality was seen in association to the use of ACEi in the context of Covid-19 disease (ACEi users versus non-users; random effects; odds ratio, 1.48; 95% confidence interval [CI], 1.02 to 2.15; P=0.04; Figure 2).

**Figure 2.**
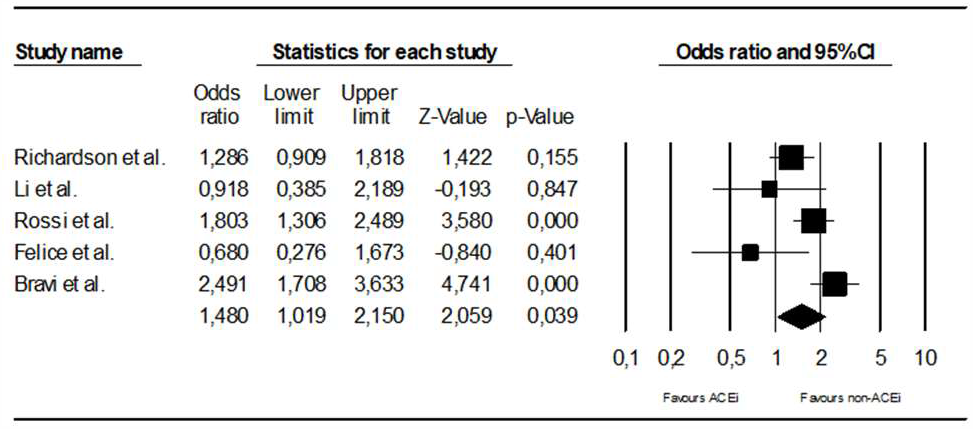
Meta-analysis comparing mortality in patients with Covid-19 disease treated or not treated with angiotensin converting inhibitors (ACEi). For references see text. CI – confidence interval.

**Figure 3.**
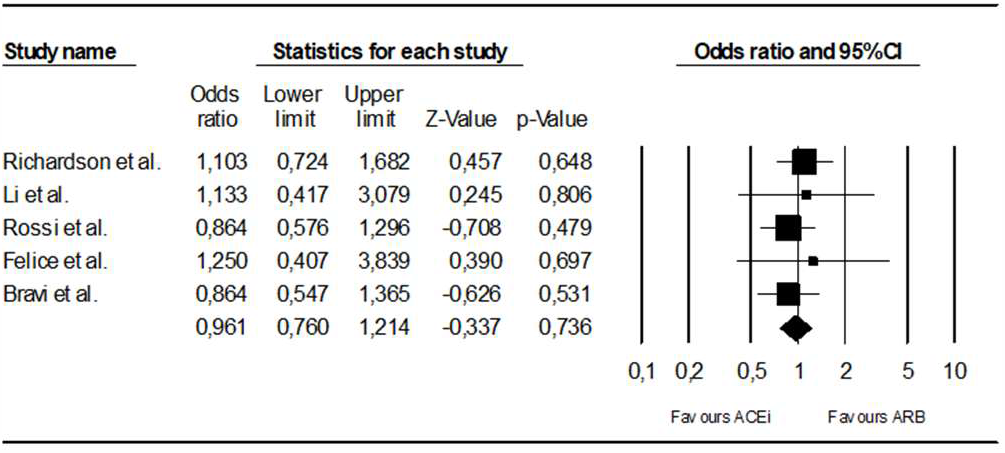
Meta-analysis comparing mortality in patients under angiotensin converting inhibitors (ACEi) or angiotensin receptor blockers (ARB) in patients with Covid-19 disease. For references see text. CI – confidence interval.

When compared to mortality in patients treated with angiotensin receptor blockers, mortality of patients treated with ACEi was not significantly different, (fixed effects, odds ratio, 0.96; 95% confidence interval [CI], 0.76 to 1.21; P=0.74) (Figure 3).

The population studied in the selected reports had different mean or median patient ages (Table 1). Overall mortality also differed when the selected reports were compared, with mortality rates ranging from under 10% to over 20% (Table 1).

The five reports used for the meta-analyses were evaluated for global quality, and the results are presented in the Supplementary Table 2 ^2^.

Rossi *et al*. indicate a numerical increase in mortality with previous ACEi use, although the authors carried out a data adjustment for age, sex and Charlson Comorbidity Index, and state that “previous use of ACE inhibitors has no effect on risk of death (HR 0.97, 95% CI 0.69 to 1.34)” ^6^. Richardson *et a*l. also show a numerical increase in mortality with ACEi use ^5^. Bravi *et al*. stated that “In multivariable analyses restricted to hypertensive subjects…, the treatment with ARBs and/or ACE inhibitors never increased the likelihood of severe or very severe/lethal disease” ^9^.

### Reports not used for the meta-analyses

A number of reports presented data on ACEi and/or ARB use in Covid-19 patients but did not contain data that could be used for the meta-analyses (listed in Supplementary Table 1; some of which presented below).

Feng *et al*. reported on 476 patients from China, and stated that ”more patients were taking angiotensin converting enzyme inhibitors/ angiotensin II receptor blockers in the moderate group than in the severe and critical groups” ^10^. Huang *et al*. reported on 50 hospitalized hypertension patients, and stated that there was no significant difference in clinical severity, clinical course and in-hospital mortality between patients either taking or not taking renin-angiotensin system blocking drugs ^11^. Zhang *et a*l. reported on 1128 adult patients with hypertension, and stated that “inpatient use of ACEI/ARB was associated with lower risk of all-cause mortality compared with ACEI/ARB nonusers” ^12^. These authors used adjusted data, ARB users were in greater number (157 patients) than ACEi users (31 patients) and 34% of patients with hypertension did not receive antihypertensive drugs during hospitalization ^12^. Zhou *et a*l. reported on a lower death rate in association to in-hospital use of ACEi or ARB therapy in COVID-19 patients with hypertension, coronary artery disease, or both ^13^.

Bean *et al*. reported on 1200 patients from the United Kingdom, and 399 Covid-19 patients were taking ACEi or ARB ^14^. The primary endpoint of death or transfer to a critical care unit was reached less often in this latter group (adjusted data).

Mancia *et a*l. reported on 6272 Covid-19 patients from Italy, as well as on a control population ^15^. The authors showed that both ARB and ACEi were more frequently prescribed in case patients than in controls. After adjustment, ARB and ACEi had no significant association with the risk of Covid-19 disease.

The report by Ip *et a*l. described favorable results for ACEi in the context of Covid-19, and the authors stated that mortality rates were lower for hypertensive patients prescribed ACEi ^16^. Argenziano *et a*l. reported on 1000 American Covid-19 patients. Hypertension was seen in 60.1% of patients, and 28.4% of patients were taking either ACEi or ARB. Reynolds *et a*l. reported on 5894 Covid-19 patients, including 2573 hypertensive patients ^17^. The authors found no association between medication class and either an increased likelihood of a positive test or of severe illness.

Khera *et a*l. studied both an outpatient and an inpatient cohort of hypertensive patients with Covid-19 disease, based on administrative data ^18^. The use of ACEi was not associated to an increased mortality risk ^18^.

## Discussion

In the present report, a systematic review was carried out, concerning the use of ACEi and a possible association to a change in mortality in Covid-19 disease. Only observational reports were found, with no clinical trial data.

In the meta-analysis, ACEi use was associated to increased mortality in the setting of Covid-19 disease. ACEi use could act as a marker of increased mortality risk in patients with Covid-19 disease – even if not causally related. ACEi use could act as a proxy for the presence of arterial hypertension, and perhaps also for the presence, in patients with arterial hypertension, of further medical conditions with an increased mortality risk – such as heart failure, chronic kidney disease, coronary artery disease or atrial fibrillation ^19^.

Arterial hypertension has been shown to act as a marker of increased mortality risk in Covid-19 disease ^20^. The age difference seen in the populations studied by the various authors could modulate the effects of ACEi use in this context. Aging is associated to an increased low grade chronic inflammatory state ^21^, decreased muscle mass, increased adiposity and a state of immune dysregulation. In Covid-19 disease, an increase in inflammatory mediators is seen in patients with more severe disease ^22^. Increasing age is a known major factor for mortality in Covid-19 disease^22^.

A number of other reports under analysis failed to show increased mortality associated to ACEi use in the setting of Covid-19 disease, after different types of statistical manipulation of data, and there were even reports presenting favorable results ^13,16^.

Should patients discontinue ACE inhibitor therapy out of a concern that they are at increased risk during the Covid-19 pandemic ^23^ ? The evidence currently available does not allow an answer to be given on a firm ground - neither a negative nor a positive one. These drugs may act as markers of increased risk in some but not all Covid-19 disease settings, but that does not mean causality exits. Until clinical trial data are available ^23^, we may consider disclosing to each patient that a high degree of uncertainty exists on this topic, and recognizing that some kind of interaction between the drug and the infectious disease process may exist, namely via the ACE2 molecule. Obtaining informed consent from the part of the patient in order to write a prescription ^24,25^ is obviously important in this context. On this matter, it is the interest and personal preferences of each person that must be taken into consideration, not the interests of society or of science, let alone those of the pharmaceutical industry.

## Limitations

Limitations of the present report are very important. Not only are all the data reviewed of an observational character, but significant differences exist when the different reports are compared, both in reports entering the meta-analyses (as shown in Table 1) and in the remaining reports under evaluation. Different types of statistical manipulation of data were used by different authors. Several types of bias could exist in the reports under study.

Significant differences exist between mortality rates associated to Covid-19 disease in different countries. The same happens with patterns of anti-hypertensive drug use.

## Conclusions

In conclusion, ACEi use could act as a marker of increased mortality risk in in some but not all Covid-19 disease settings. The data now presented do not prove a causal relation but argue in favor of carrying out clinical trials studying ACEi in Covid-19 patients, in order to establish the safety of ACEi use in this context.

## Data Availability

All data was obtained from the cited articles

## References

1. Hoffmann M, Kleine-Weber H, Schroeder S, et al. SARS-CoV-2 Cell Entry Depends on ACE2 and TMPRSS2 and Is Blocked by a Clinically Proven Protease Inhibitor. Cell 2020;181:271-80.e8.

2. Haffar S, Shalimar Kaur RJ, et al. Acute liver failure caused by hepatitis E virus genotype 3 and 4: A systematic review and pooled analysis. Liver Int 2018;38:1965–73.

3. Meng J, Xiao G, Zhang J, et al. Renin-angiotensin system inhibitors improve the clinical outcomes of COVID-19 patients with hypertension. Emerging Microbes & Infections 2020;9:757–60.

4. Jung S-Y, Choi JC, You S-H, Kim W-Y. Association of renin-angiotensin-aldosterone system inhibitors with COVID-19-related outcomes in Korea: a nationwide population-based cohort study. Clinical Infectious Diseases 2020.

5. Richardson S, Hirsch JS, Narasimhan M, et al. Presenting Characteristics, Comorbidities, and Outcomes Among 5700 Patients Hospitalized With COVID-19 in the New York City Area. JAMA 2020.

6. Giorgi Rossi P, Marino M, Formisano D, Venturelli F, Vicentini M, Grilli R. Characteristics and outcomes of a cohort of SARS-CoV-2 patients in the Province of Reggio Emilia, Italy. medRxiv 2020:2020.04.13.20063545.

7. Li J, Wang X, Chen J, Zhang H, Deng A. Association of Renin-Angiotensin System Inhibitors With Severity or Risk of Death in Patients With Hypertension Hospitalized for Coronavirus Disease 2019 (COVID-19) Infection in Wuhan, China. JAMA Cardiology 2020.

8. Felice C, Nardin C, Di Tanna GL, et al. Use of RAAS inhibitors and risk of clinical deterioration in COVID-19: results from an Italian cohort of 133 hypertensives. American Journal of Hypertension 2020.

9. Bravi F, Flacco ME, Carradori T, et al. Predictors of severe or lethal COVID-19, including Angiotensin Converting Enzyme inhibitors and Angiotensin II Receptor Blockers, in a sample of infected Italian citizens. PLoS One 2020;15:e0235248.

10. Feng Y, Ling Y, Bai T, et al. COVID-19 with Different Severities: A Multicenter Study of Clinical Features. Am J Respir Crit Care Med 2020;201:1380–8.

11. Huang Z, Cao J, Yao Y, et al. The effect of RAS blockers on the clinical characteristics of COVID-19 patients with hypertension. Ann Transl Med 2020;8:430.

12. Zhang P, Zhu L, Cai J, et al. Association of Inpatient Use of Angiotensin-Converting Enzyme Inhibitors and Angiotensin II Receptor Blockers With Mortality Among Patients With Hypertension Hospitalized With COVID-19. Circ Res 2020;126:1671–81.

13. Zhou F, Liu Y-M, Xie J, et al. Comparative impacts of angiotensin converting enzyme inhibitors versus angiotensin II receptor blockers on the risk of COVID-19 mortality. Hypertension;0.

14. Bean DM, Kraljevic Z, Searle T, et al. ACE-inhibitors and Angiotensin-2 Receptor Blockers are not associated with severe SARS-COVID19 infection in a multi-site UK acute Hospital Trust. Eur J Heart Fail 2020.

15. Mancia G, Rea F, Ludergnani M, Apolone G, Corrao G. Renin–Angiotensin–Aldosterone System Blockers and the Risk of Covid-19. New England Journal of Medicine 2020.

16. Ip A, Parikh K, Parrillo JE, et al. Hypertension and Renin-Angiotensin-Aldosterone System Inhibitors in Patients with Covid-19. medRxiv 2020:2020.04.24.20077388.

17. Reynolds HR, Adhikari S, Pulgarin C, et al. Renin–Angiotensin–Aldosterone System Inhibitors and Risk of Covid-19. New England Journal of Medicine 2020.

18. Khera R, Clark C, Lu Y, et al. Association of Angiotensin-Converting Enzyme Inhibitors and Angiotensin Receptor Blockers with the Risk of Hospitalization and Death in Hypertensive Patients with Coronavirus Disease-19. medRxiv 2020:2020.05.17.20104943.

19. Williams B, Mancia G, Spiering W, et al. 2018 ESC/ESH Guidelines for the management of arterial hypertension: The Task Force for the management of arterial hypertension of the European Society of Cardiology (ESC) and the European Society of Hypertension (ESH). European Heart Journal 2018;39:3021–104.

20. Cummings MJ, Baldwin MR, Abrams D, et al. Epidemiology, clinical course, and outcomes of critically ill adults with COVID-19 in New York City: a prospective cohort study. Lancet 2020.

21. Chung HY, Kim DH, Lee EK, et al. Redefining Chronic Inflammation in Aging and Age-Related Diseases: Proposal of the Senoinflammation Concept. Aging Dis 2019;10:367–82.

22. Huang C, Wang Y, Li X, et al. Clinical features of patients infected with 2019 novel coronavirus in Wuhan, China. Lancet 2020;395:497–506.

23. Jarcho JA, Ingelfinger JR, Hamel MB, D’Agostino RB, Harrington DP. Inhibitors of the Renin–Angiotensin–Aldosterone System and Covid-19. New England Journal of Medicine 2020.

24. Nunes JPL. Medical therapeutics: mortality effects, uncertainty, and informed consent. Porto Biomedical Journal 2019;4:e35.

25. Colloca L, Barsky AJ. Placebo and Nocebo Effects. New England Journal of Medicine 2020;382:554–61.

